# The impact of post-traumatic stress disorder in pharmacological intervention outcomes for adults with bipolar disorder: a protocol for a systematic review and meta-analysis

**DOI:** 10.1101/2022.05.02.22274560

**Authors:** Samantha E. Russell, Anna L. Wrobel, David Skvarc, Bianca E Kavanagh, Melanie M Ashton, Olivia M. Dean, Michael Berk, Alyna Turner

## Abstract

**Background:** Recent data indicates high prevalence of post-traumatic stress disorder (PTSD) in bipolar disorder (BD). PTSD may play a role in poor treatment outcomes and quality of life for people with BD. Despite this, few studies have examined the pharmacological treatment interventions and outcomes for this comorbidity. This systematic review will bring together currently available evidence regarding the impact of comorbid PTSD on pharmacological treatment outcomes in adults with BD.

**Methods:** A systematic search of Embase, MEDLINE Complete, PsycINFO, and the Cochrane Central Register of Controlled Trials (CENTRAL) will be conducted to identify randomised and non-randomised studies of pharmacological interventions for adults with diagnosed bipolar disorder and PTSD. Data will be screened and extracted by two independent reviewers. Literature will be searched from the creation of the databases until April 1 2021. Risk of bias will be assessed using the Newcastle-Ottawa Scale and the Cochrane Collaborations Risk of Bias tool. A meta-analysis will be conducted if sufficient evidence is identified in the systematic review. The meta-analysis will employ a random-effects model and be evaluated using the I^2^ statistic.

**Discussion:** This review and meta-analysis will be the first to systematically explore and integrate the available evidence on the impact of PTSD on pharmacological treatments and outcome in those with BD. The results and outcomes of this systematic review will provide directions for future research and be published in relevant scientific journals and presented at research conferences.

**Systematic review registration:** The protocol has been registered at the International Prospective Register of Systematic Reviews (PROSPERO; registration number: CRD42020182540).

## Background

Evidence suggests that Post-traumatic Stress Disorder (PTSD) and bipolar disorder (BD) commonly co-occur. Rates of PTSD have been found to be much higher in people with BD than in the general population,[1] with estimates varying from 7% to 55%.[2] for the comorbidity. Bipolar disorder is among the top 20 leading causes of disability worldwide according to the Global Disease Burden Study in 2013.[3] These two psychiatric disorders together create a high-risk and vulnerable population with an increased mental and physical health and quality of life burden.[4–6] Crucially, the overlap in symptomatology in PTSD and BD may lead to underdiagnosis or misdiagnosis.[1] The disorders share some common symptoms and features including sleep disruptions, difficulty concentrating, irritability, difficulty maintaining relationships or employment, anhedonia, mood swings, anxiety and hopelessness.[1] Furthermore, it can be difficult to differentiate psychomotor agitation from a hypomanic/manic episode.[1] Emotional lability induced by environmental triggers (e.g. sudden panic, distress and avoidance) experienced in PTSD is associated with risk for relapse for BD episodes.[7]

PTSD has an impact on BD.[8] Cross-sectional analysis of hospitalised US veterans,[8] primary care[6] and a sample of Brazilian patients with Bipolar Disorder 1 (BDI)[9] found those with comorbid BD and PTSD experience more severe symptoms, increased rapid cycling, more manic episodes, lower functioning scores, higher rates of disability pension use, worse quality of life, and more disability than those with BD alone. A European cross-sectional sample[4] and a retrospective chart review of BDI patients[10] found those with comorbid BD and PTSD were more vulnerable and had higher risks of exposure to physical violence, alcoholism and sexual assault, spent more time depressed and had a history of more complex polypharmacy compared to those with BD alone.

Recent studies by Carter and colleagues (2017)[11] and Katz and colleagues (2020)[5] investigated the increased suicide risk in those with comorbid BD and PTSD. Both cross-sectional analyses found increased suicidal ideation, suicide attempts and lower quality of life compared those with BD alone. In addition, Quarantini and colleagues[9] emphasise the need to assess BD patients for PTSD as it is a predictor of suicide risk. Most relevant to the current review, these studies found that patients with comorbid BD and PTSD had a lower likelihood of staying recovered, increased suicide attempts, and increased rates of illness severity.[4]

While psychotherapy remains the foundation approach, pharmacological treatment strategies for PTSD as recommended by The Australian Guidelines for the Treatment of Acute Stress Disorder & Posttraumatic Stress Disorder[12] involve mono- or combination therapies of antidepressants, antipsychotics, mood stabilisers, anti-convulsants, and beta-blockers. Selective serotonin reuptake inhibitors (SSRIs) escitalopram, fluoxetine and paroxetine have clinical evidence to support the use in PTSD. Similarly, the National Institute for Health and Care Excellence (NICE) guidelines[13] recommend the use of SSRI’s (particularly venlafaxine, paroxetine or sertraline) and antipsychotic risperidone (if disabling symptoms are present such as psychosis and these symptoms have not responded to other treatments) for treatment of PTSD. The Australian Guidelines also recommend the use of adjunctive antipsychotics (risperidone, olanzapine, clozapine and quetiapine) for complex or treatment-resistant cases.[12] Additionally, benzodiazepines are occasionally used to assist with anxiety and insomnia.[12] However, new WHO recommendations warn against the use of benzodiazepines in PTSD stating there is no evidence on the benefits of benzodiazepines on symptoms of traumatic stress, and they may slow down recovery time from the traumatic event.[14] Moreover, WHO Guidelines for the management of conditions related explicitly to stress[14] recommend against the use of SSRIs and tricyclic antidepressants as the first line of treatment in PTSD. The WHO recommends their use only if psychotherapies have failed or there is comorbid moderate to severe depression.

Pharmacotherapies for BD consist of mood stabilisers, anti-convulsants, anxiolytics and antipsychotics.[15],[16] There is an unmet need for clinical trials evaluating the effectiveness of pharmacological treatments in comorbid BD and PTSD, especially as antidepressants are also the first-line treatment in PTSD.[5] A meta-analysis investigating the prevalence of antidepressant-induced mania in BD found a high incidence (pooled prevalence of 30.9%) of manic-switch and caution against their use in BD as a mono-therapy.[17] There is an inconsistency between the guideline-recommended pharmacological treatment options for both BD and PTSD. Pharmacological options for PTSD recommended by the WHO guidelines recommend against benzodiazepines and anti-depressants for PTSD, whereas The Australian Guidelines and The NICE guidelines[13] recommend for anti-depressant use and benzodiazepine use if needed. Additionally, the risk of manic switch in BD from anti-depressants prescribed for PTSD creates further complications. This misalliance of guideline recommendations may be an important confounding factor in the pharmacological treatment of comorbid BD and PTSD.

Despite the overlap in symptoms, high prevalence rates and increased illness burden of those comorbid BD and PTSD, few studies have assessed and/or evaluated prospective treatment strategies in people with comorbid BD and PTSD.[7] A recent rapid review[2] on treatments for this comorbidity found no trials that have evaluated pharmacological treatments for this population. This systematic review will build upon the 2017 rapid review, which largely focussed on epidemiology, clinical correlates and prevalence estimates of comorbid BD and PTSD. For example, a meta-analysis is planned in this systematic review and it will focus on outcomes of pharmacological treatments. The identification of effective pharmacological treatments for this comorbidity is vitally important to help reduce the increased burden of illness. Previous research has suggested that comorbid PTSD influences treatment outcomes in bipolar disorder. Pharmacogenomic and cohort studies have found people with comorbid BD and PTSD have a decreased response to lithium compared to those BD alone.[18],[19] Interestingly, the Fornaro[17] meta-analysis suggests that lithium is among the most effective treatments for treatment-resistant mania. Previous research by Nierenberg[20] and Carter and collegues[11] have identified the challenges of pharmacotherapy amongst this comorbidity by highlighting that those with comorbid PTSD and BD show poorer response to medication and are less likely to adhere to treatment plans.[11,20]

Due to the complexity of the clinical presentation of comorbid BD and PTSD, polypharmacy is often employed.[10],[21] Given the lack of information and inconsistent findings concerning pharmacological interventions and treatment outcomes in comorbid BD and PTSD, this systematic review is warranted. The proposed review will bring together available evidence regarding pharmacological treatments and outcomes for people with comorbid BD and PTSD, contrasted to those with BD alone, to inform treatment decision making for clinicians and patients. The focus of this review is on exploring outcomes for people with bipolar disorder with and without comorbid PTSD.

### Objectives

The aims of this systematic review are:

1. To identify and synthesise published studies of pharmacological interventions and treatment outcomes within a population of comorbid BD and PTSD and BD alone. This includes studies of BD, which also include a PTSD comorbid cohort and assessment.
2. To appraise the quality of the methodology used in each study eligible for inclusion in this systematic review. Specifically, to provide a synthesis and meta-analysis of the evidence and to evaluate whether treatment outcomes differ in those with comorbid BD and PTSD and those with BD alone.

### Methods and analysis

The PICO framework (Populations, Intervention, Comparator and Outcome) was used to develop the search strategy. We used the Preferred Reporting Items for Systematic Review and Meta-Analysis Protocols PRISMA-P checklist when writing this protocol see additional file 1 for more details.[22]

#### Participants/population

Adults (18 years or over) diagnosed with BDI, BDII or other bipolar subtypes and diagnosed with or without PTSD will be included in the review. If studies include participants under 18 years of age and 18 years of age or older, only the participant data of those 18 years of age or older (if available) will be included in the systematic review. Diagnosis of BD must be confirmed by diagnostic interview, either via a structured or semi-structured clinical interview (e.g. The Composite International Diagnostic Interview [CIDI],[23] The Structured Clinical Interview for the DSM [SCID][24] The Mini International Neuropsychiatric Interview [MINI][25]) or via psychiatrist. Diagnosis of PTSD must be validated via screening tool (e.g. Clinician administered PTSD Scale [CAPS-5][26], Posttraumatic Diagnostic Scale [PDS][27], PTSD Checklist for DSM-5 [PCL-5][28]), psychiatrist diagnosis or structured or semi-structured clinical interview, such as; any version of CIDI,[23] SCID,[24] or MINI. Studies are to include people with a diagnosis of BD and an assessment or diagnosis of PTSD. If studies have reported that PTSD assessment was undertaken but not reported in the analysis, or missing data is identified, attempts will be made to contact the authors to obtain the data.

#### Intervention

All pharmacological treatments for symptoms of BD and PTSD will be reviewed. Treatments may include but are not limited to mood stabilisers, antidepressants and antipsychotics.

#### Comparator or control

Any intervention, non-exposed control groups, waitlist controls, active comparators, placebo, treatment as usual or standard care comparisons or controls are to be included.

#### Types of study to be included

Studies to be included are randomised trials, non-randomised trials, cross-over trials, randomised controlled trials (RCT), cluster RCTs, one arm trials, controlled non randomised trials, cohort studies, case-control studies, cross-sectional studies and open-label studies. Studies in community, clinical and hospital settings will be included, in all social-economic statuses and all countries. Studies to be excluded are case reports, case series and qualitative studies.

### Search strategy

Medical, health and psychology databases will be electronically searched. These consist of EMBASE via embase.com, PsycINFO via EBSCO, CENTRAL via cochranelibrary.com, and Medline via EBSCO. Additionally, citation searching will be completed using Scopus via elsevier.com before and after the searches to ensure all relevant articles have been captured. The following medical subject headings (MeSH) and search terms will be used: (‘mood disorders’ OR ‘mood disorder’ OR ‘bipolar and related disorders’ OR ‘affective disorders’ OR ‘bipolar disorder’ OR ‘mania’ OR ‘manic depression’ OR ‘bipolar depression’ OR ‘severe mental illness’ OR ‘serious mental illness’ OR ‘SMI’) AND (‘PTSD’ OR ‘post-traumatic stress disorders’ OR ‘post-traumatic stress disorder’ OR ‘post-traumatic stress disorder’ OR ‘post-traumatic stress disorder’) AND (‘drug therapy’ OR ‘pharmacotherapy’ OR ‘pharmaceuticals’ OR ‘intervention’ OR ‘drugs’ OR ‘pharmacology’ OR ‘medication” OR ‘trial’ OR ‘antipsychotics’ OR ‘mood stabiliser’ OR ‘antidepressant’ OR ‘anxiolytics’ OR ‘hypnosedatives’). Appropriate wildcard symbols and truncation will be applied in each database. Literature will be searched from the creation of the databases until April 1 2021. A preliminary search strategy is displayed in Additional File 2.

The reference lists of articles included in the review will also be hand searched for any relevant articles not found in the electronic database search. All selected studies will be downloaded into Mendeley, and duplicates will be removed. Papers in languages other than English will be excluded. The authors of the original studies will be contacted for additional information if relevant outcomes of interest are not reported. Searches will be rerun prior to the final analysis to identify any further studies. Unpublished studies will not be sought for this review.

### Main outcome

The primary outcome will be to evaluate the impact of PTSD on treatment outcomes in studies of pharmacological interventions for adults with bipolar disorder. Specifically, to measure any change in BD outcome score from baseline to the end of the study period. Treatment outcomes will be measured with any validated assessment tool, such as Montgomery Åsberg Rating Scale (MADRS),[29] Bipolar Depression Rating Scale (BDRS)[30], Young Mania Rating Scale (YMRS),[31] and Bipolar Inventory of Symptom Scale (BISS)[32] and any other validated scales assessing bipolar disorder will also be considered.

#### Secondary outcomes

Secondary outcomes of this systematic review include assessing the impact that PTSD has on participants’ subjective outcomes, such as self-reported symptom improvements and assessments of the quality of life and functioning (e.g., the Quality of Life Enjoyment and Satisfaction Questionnaire [Q-LES-Q][33], Clinical Global Impressions-Bipolar Disorder (GCI-BP)[34], Clinical Global Impressions – Improvement (CGI-I score),[34] clinician-rated functioning scales: Social and Occupational Functioning Assessment Scale [SOFAS][35], Longitudinal Interval Follow-up Evaluation – Range of Impaired Functioning Tool [LIFE-RIFT][36] and any other validated scale assessing functioning or quality of life).

### Data extraction (selection and coding)

Titles and abstracts identified in the searches will be screened by two authors to determine whether they are eligible for inclusion. Full-text articles will be retrieved from studies that satisfy the eligibility criteria of pharmacological treatment in adult participants with a diagnosis of BD and an assessment of PTSD or a comorbid diagnosis of BD and PTSD. Full texts will then be read and assessed for eligibility by two authors, with disagreements resolved by discussion until consensus is reached or by utilising a third author. The Covidence platform will be used for screening and selection of studies for review, and a custom REDCap[37,38] extraction tool will be utilised

Two authors will independently extract the data. Extracted data will include:

1. Study characteristics for identification (author name, publication year, country or countries of study)
2. Study design (e.g. randomised, cross-sectional, longitudinal)
3. Sample size and characteristics
5. BD type (I, II or subtype)
6. Details of pharmacological treatments or interventions
7. Demographic characteristics
8. Diagnostic and assessment tools utilised
9. Study outcomes and results (main and secondary outcomes)

#### Evaluation of methodological quality of included articles

Cochrane Collaborations Risk of Bias tool[39] will be utilised for randomised controlled trials, and eligible literature will be scored low, high or unclear risk of bias. Specifically, the domains of bias include selection bias, performance bias, detection bias, attrition bias, reporting bias and any other bias detected. For non-randomised studies, the Newcastle-Ottawa Scale[40] will be used. The Joanna Briggs Institute (JBI) critical appraisal tools[41] will also be utilised if other observational study designs are identified for the review. The Newcastle Ottawa scale grades on selection, comparability and outcomes using a star award system. The quality of studies could be defined as high (10-9 stars), moderate (7-8 stars), low (6 stars and below).[40]

#### Strategy for data synthesis

Data will be analysed for quality of evidence using the Grades of Recommendation, Assessment, Development and Evaluation (GRADE) procedure.[42] The quality of evidence will be graded as high, moderate, low and very low. Limitations in comprehensive design, heterogeneity or inconsistency, indirectness, imprecision and publication bias will also be assessed. The GRADE[42] will analyse data for quality of evidence as per guidelines of the JBI[41] whether a meta-analysis can or cannot be conducted.

A meta-analysis will be conducted if more than two eligible studies are identified. Randomised and non-randomised intervention studies will be analysed and presented separately. The meta-analysis will involve a random-effects model, and 95% confidence intervals (CI) and a p-value will be reported. This meta-analysis requires that the treatment effects have been reported in the studies according to BD subgroup (i.e. BD+PTSD vs BD only) and the mean symptom change of BD between participants with and without comorbid PTSD. For continuous data, the standard mean difference (SMD) will be calculated with 95% CIs. Where possible, differences in treatment outcomes between disorder groups will be compared using mean differences on the same rating scale. Where different outcomes measures are used, standardized mean difference will be used. Effect sizes will be calculated using Hedges’ g. Sensitivity analysis will be conducted to determine the robustness of the meta-analysis outcomes. For example, studies with a high rating on the New Castle Ottawa and those with a moderate and high rating will be compared; however, other sensitivity analysis may be performed.

The meta-analysis will be conducted with the Comprehensive Meta-Analysis (CMA) software[43] and heterogeneity of evidence will be determined using the Higgins I^2^ statistic calculations.[44] If substantial heterogeneity between studies is found (I^2^>50%), the possible reasons for between-study variability will be considered by analysing the included studies characteristics, such as the methodological differences (e.g., outcome measures used) and sources of potential bias will be explored. A random-effects model will be utilised if a meta-analysis is possible. If a meta-analysis is not possible, a narrative synthesis will be conducted.

### Presentation and reporting of results

We will use the Preferred Reporting Items for Systematic Review and Meta-Analysis PRISMA checklist when writing our report. The result of the searches and screening process will be reported in a PRISMA flow diagram, including the number of studies and reasons regarding included versus excluded studies at each stage.

## Discussion

This planned review and meta-analysis will systematically explore the available evidence on pharmacological treatments in adults with BD+PTSD. This review will aim to compare pharmacological treatments and associated outcomes between those with BD only, and those with PTSD+BD. The findings from this study will provide directions for future research and provide clinicians with an understanding of the current treatment landscape for those with BD+PTSD. This knowledge can be used to develop more informed treatment strategies and interventions.

## Supporting information

Additonal File 1.

Additonal File 2.

## Data Availability

This is a systematic review protocol and therefore no data is available.

## List of abbreviations

BD: bipolar disorder
PTSD: post-traumatic stress disorder
CENTRAL: Cochrane Central Register of Controlled Trials
BDI: Bipolar Disorder 1
SSRI: Selective serotonin reuptake inhibitors
NICE: National Institute for Health and Care Excellence
WHO: World Health Organization
PICO: Populations, Intervention, Comparator and Outcome
PRISMA: P Preferred Reporting Items for Systematic Review and Meta-Analysis Protocols
CIDI: The Composite International Diagnostic Interview
SCID: Structured Clinical Interview for the DSM
MINI: The Mini International Neuropsychiatric Interview
CAPS: Clinician administered PTSD Scale
PDS: Posttraumatic Diagnostic Scale
PCL5: PTSD Checklist for DSM-5
RCT: Randomised controlled trials
MeSH: Medical subject headings
MADRS: Montgomery Åsberg Rating Scale
BDRS: Bipolar Depression Rating Scale
YMRS: Young Mania Rating Scale
BISS: Bipolar Inventory of Symptom Scale
Q-LES-Q: Quality of Life Enjoyment and Satisfaction Questionnaire
CGI-BP: Clinical Global Impressions-Bipolar Disorder
CGI-I: Clinical Global Impressions – Improvement
SOFAS: Social and Occupational Functioning Assessment Scale
LIFE-RIFT: Longitudinal Interval Follow-up Evaluation – Range of Impaired Functioning Tool
GRADE: the Grades of Recommendation, Assessment, Development and Evaluation
JBI: Joanna Briggs Institute
CI: Confidence intervals
SMD: Standard mean difference
CMA: Comprehensive Meta-Analysis

## Declarations

### Ethics approval and consent to participate

Not applicable. Ethical governance principles will be complied with regarding data management and the dissemination/presentation of findings.

### Consent for publication

Not applicable.

### Availability of data and materials

The studies included in the review will be available upon request.

### Author Contributions

**SER, AT** and **OMD** conceptualised and designed the research question and developed the search strategy. **SER, ALW, DS, MMA, BEK, OMD, MB** and **AT** edited, revised and approved the final version of the manuscript.

## Acknowledgements

The authors would like to acknowledge and thank Medical Librarian Blair Kelly, Deakin University for assisting with the search strategy design and development.

## Additional Files

Additional File 1 in word document format (*.docx). PRISMA-P 2015 Checklist.

Additional File 2 in word document format (*.docx). MEDLINE Complete preliminary search strategy.

## References

1. Hernandez JM, Cordova MJ, Ruzek J, Reiser R, Gwizdowski IS, Suppes T, et al. Presentation and prevalence of PTSD in a bipolar disorder population: A STEP-BD examination. J Affect Disord. 2013;

2. Cerimele JM, Bauer AM, Fortney JC, Bauer MS. Patients with co-occurring bipolar disorder and posttraumatic stress disorder: A rapid review of the literature. J. Clin. Psychiatry. Physicians Postgraduate Press Inc.; 2017. p. e506–14.

3. Ferrari AJ, Stockings E, Khoo JP, Erskine HE, Degenhardt L, Vos T, et al. The prevalence and burden of bipolar disorder: findings from the Global Burden of Disease Study 2013. Bipolar Disord. 2016;18:440–50.

4. Assion HJ, Brune N, Schmidt N, Aubel T, Edel MA, Basilowski M, et al. Trauma exposure and post-traumatic stress disorder in bipolar disorder. Soc Psychiatry Psychiatr Epidemiol. 2009;44:1041–9.

5. Katz D, Petersen T, Amado S, Kuperberg M, Dufour S, Rakhilin M, et al. An evaluation of suicidal risk in bipolar patients with comorbid posttraumatic stress disorder. J Affect Disord. Elsevier B.V.; 2020;266:49–56.

6. Neria Y, Olfson M, Gameroff MJ, Wickramaratne P, Pilowsky D, Verdeli H, et al. Trauma exposure and posttraumatic stress disorder among primary care patients with bipolar spectrum disorder. Bipolar Disord. 2008;10:503–10.

7. Otto MW, Perlman CA, Wernicke R, Reese HE, Bauer MS, Pollack MH. Posttraumatic stress disorder in patients with bipolar disorder: a review of prevalence, correlates, and treatment strategies. Bipolar Disord. 2004.

8. Bauer MS, Altshuler L, Evans DR, Beresford T, Williford WO, Hauger R. Prevalence and distinct correlates of anxiety, substance, and combined comorbidity in a multi-site public sector sample with bipolar disorder. J Affect Disord. 2005;85:301–15.

9. Quarantini LC, Miranda-Scippa Â, Nery-Fernandes F, Andrade-Nascimento M, Galvão-de-Almeida A, Guimarães JL, et al. The impact of comorbid posttraumatic stress disorder on bipolar disorder patients. J Affect Disord. 2010;123:71–6.

10. Reddy MK, Meyer TD, Wittlin NM, Miller IW, Weinstock LM. Bipolar I disorder with comorbid PTSD: Demographic and clinical correlates in a sample of hospitalized patients. Compr Psychiatry. 2017;72:13–7.

11. Carter JM, Arentsen TJ, Cordova MJ, Ruzek J, Reiser R, Suppes T, et al. Increased Suicidal Ideation in Patients with Co-Occurring Bipolar Disorder and Post-Traumatic Stress Disorder. Arch Suicide Res. Routledge; 2017;21:621–32.

12. Phoenix Australia - Centre for Posttraumatic Mental Health. Australian Guidelines for the Treatment of Acute Stress Disorder and Posttraumatic Stress Disorder. Phoenix Aust. Melb. Vic. 2013.

13. National Institute for Health and Care Excellence. Post-traumatic Stress disorder NICE guideline [NG116]. 2018. p. 21–3.

14. World Health Organization. Guidelines for the management of conditions specifically related to stress. Geneva; 2013.

15. Gin S Malhi, Darryl Bassett PB, Richard Bryant, Paul B Fitzgerald KF, Malcolm Hopwood BL, Roger Mulder, Greg Murray RP, Singh and AB. Royal Australian and New Zealand College of Psychiatrists clinical practice guidelines for mood disorders: Major depression summary. Aust N Z J Psychiatry. 2015;49:1–185.

16. National Institute for Health and Care Excellence. Bipolar disorder: assessment and management. Aust Fam Physician. 2020;36:12–9.

17. Fornaro M, Anastasia A, Novello S, Fusco A, Solmi M, Monaco F, et al. Incidence, prevalence and clinical correlates of antidepressant-emergent mania in bipolar depression: a systematic review and meta-analysis. Bipolar Disord. 2018;20:195–227.

18. Bremer T, Diamond C, McKinney R, Shehktman T, Barrett TB, Herold C, et al. The pharmacogenetics of lithium response depends upon clinical co-morbidity. Mol Diagn Ther. 2007;11:161–70.

19. Cakir S, Tasdelen Durak R, Ozyildirim I, Ince E, Sar V. Childhood trauma and treatment outcome in bipolar disorder. J Trauma Dissociation. 2016;17:397–409.

20. Nierenberg AA. Lessons from STEP-BD for the treatment of bipolar depression. Depress Anxiety. 2009;26:106–9.

21. Golden JC, Goethe JW, Woolley SB. Complex psychotropic polypharmacy in bipolar disorder across varying mood polarities: A prospective cohort study of 2712 inpatients. J Affect Disord. 2017;221:6–10.

22. Moher D, Shamseer L, Clarke M, Ghersi D, Liberati A, Petticrew M, Shekelle P S LA. Preferred Reporting Items for Systematic Review and Meta-Analysis Protocols (PRISMA-P) 2015 statement. Syst Rev. 2015;4.

23. Robins LN. The Composite International Diagnostic Interview (CIDI). 1994;250–4.

24. First, BM. Structured Clinical Interview for DSM-IV Axis I Disorders. Biom Res Dep. 1997;

25. Lecrubier Y, Sheehan DV, Weiller E, Amorim P, Bonora I, Sheehan KH, et al. The Mini International Neuropsychiatric Interview (MINI). A short diagnostic structured interview: Reliability and validity according to the CIDI. Eur Psychiatry. Éditions scientifiques et médicales 1997;12:224–31.

26. Boeschoten MA, Van der Aa N, Bakker A, Ter Heide FJJ, Hoofwijk MC, Jongedijk RA, et al. Development and Evaluation of the Dutch Clinician-Administered PTSD Scale for DSM-5 (CAPS-5). Eur J Psychotraumatology. 2018;9.

27. Foa EB. The posttraumatic diagnostic scale (PDS) manual. Natl Comput Syst. Minneapolis: MN: National Computer Systems; 1995;

28. Blevins CA, Weathers FW, Davis MT, Witte TK, Domino JL. The Posttraumatic Stress Disorder Checklist for DSM-5 (PCL-5): Development and Initial Psychometric Evaluation. J Trauma Stress. 2015;28:489–98.

29. Montgomery SA, Asberg M. A new depression scale designed to be sensitive to change. Br J Psychiatry. 1979;134:382–89

30. Berk M, Malhi GS, Cahill C, Carman AC, Hadzi-Pavlovic D, Hawkins MT, et al. The Bipolar Depression Rating Scale (BDRS): Its development, validation and utility. Bipolar Disord. 2007;9:571–9.

31. Young RC, Biggs JT, Ziegler VE, Meyer DA. Young Mania Rating Scale. Can J Clin Pharmacol. 2004;540–42

32. Gonzalez JM, Bowden CL, Katz MM, Thompson P, Singh V, Prihoda TJ, et al. Development of the Bipolar Inventory of Symptoms Scale: concurrent validity, discriminant validity and retest reliability. Int J Methods Psychiatr Res. 2008;17:198–209.

33. Endicott J, Nee J, Harrison W, Blumenthal R. Quality of life enjoyment and satisfaction questionnaire: A new measure. Psychopharmacol Bull. 1993;29;321–26.

34. Spearing MK, Post RM, Levericha GS, Brandtb D, Nolenc W. Modification of the Clinical Global Impressions (CGI) scale for use in bipolar illness (BP): the CGI-BP. 1997;73:159–71.

35. Morosini PL, Magliano L, Brambilla L, Ugolini S, Pioli R. Development, reliability and acceptability of a new version of the DSM-IV Social and Occupational Functioning Assessment Scale (SOFAS) to assess routine social functioning. Acta Psychiatr Scand. 2000;101:323–9.

36. Keller MB. The Longitudinal Interval Follow-up Evaluation. Arch Gen Psychiatry. 1987;44:540.

37. PA Harris, R Taylor, R Thielke, J Payne, N Gonzalez JGC. Research electronic data capture (REDCap) – A metadata-driven methodology and workflow process for providing translational research informatics support. J Biomed Inf. 2009;42:377–81.

38. PA Harris, R Taylor, BL Minor, V Elliott, M Fernandez, L O’Neal, L McLeod, G Delacqua, F Delacqua, J Kirby SD. REDCap Consortium, The REDCap consortium: Building an international community of software partners. J Biomed Inf. 2019;95.

39. Higgins JPT, Altman DG, Gøtzsche PC, Jüni P, Moher D, Oxman AD. The Cochrane Collaboration ‘ s tool for assessing risk of bias in randomised trials. 2011;1–9.

40. Wells GA, Shea B, O’Connell D, Peterson J, Welch V, Losos M, et al. The Newcastle-Ottawa Scale (NOS) for Assessing the Quality of Nonrandomized Studies in Meta-Analysis. https://www.ohri.ca/programs/clinical_epidemiology/oxford.asp. Accessed 8 June 2021

41. JBI Institute. The Joanna Briggs Institute Critical Appraisal tools for use in JBI Systematic Reviews. 2020. https://jbi.global/critical-appraisal-tool. Accessed 30 May 2021.

42. Guyatt GH, Oxman AD, Vist GE, Kunz R, Falck-Ytter Y, Alonso-Coello P, et al. GRADE: An emerging consensus on rating quality of evidence and strength of recommendations. BMJ. BMJ Publishing Group; 2008;336:924–6.

43. Biostat. Comprehensive Meta-Analysis [program] 3.0 version. Engelwood: Biostat, Inc;

44. Higgins JPT, Thomas J, Chandler J, Li T, Page MJ, Welch VA. Cochrane Handbook for Systematic Reviews of Interventions version 6.2 (updated February 2021). Cochrane. 2021. http://www.training.cochrane.org/handbook. Accessed 12 July 2021.

