## Supplementary material for "The impact of post-traumatic stress disorder in pharmacological intervention outcomes for adults with bipolar disorder: a protocol for a systematic review and meta-analysis": Additonal File 2.

Additional File 2. MEDLINE Complete preliminary search strategy.

| **#** | **Medline Query Final** |
| --- | --- |
| S35 | s32 and s33 and s34 |
| S34 | s16 or s17 or s18 or s19 |
| S33 | s1 or s2 or s3 or s4 or s5 or s6 or s7 or s8 or s9 or s10 or s11 or s12 or s13 or s14 or s15 |
| S32 | s20 or s21 or s22 or s23 or s24 or s25 or s26 or s27 or s28 or s29 or s30 or s31 |
| S31 | TI ("treatment outcome*") or AB ("treatment outcome*") |
| S30 | TI ("treatment strateg*") or AB ("treatment strateg*") |
| S29 | TI (drug*) or AB (drug*) |
| S28 | TI (intervention*) or AB (intervention*) |
| S27 | TI (pharmacolog*) or AB (pharmacolog*) |
| S26 | TI (pharmaceutical*) or AB (pharmaceutical*) |
| S25 | TI (pharmacotherap*) or AB (pharmacotherap*) |
| S24 | (MH "Treatment Outcome") |
| S23 | (MH "Pharmaceutical Preparations+") |
| S22 | (MH "Clinical Trials as Topic+") |
| S21 | MH (pharmacology+) |
| S20 | MH ("drug therapy+") |
| S19 | TI ("posttraumatic stress disorder") OR AB ("posttraumatic stress disorder") |
| S18 | TI ("post traumatic stress disorder") OR AB ("post traumatic stress disorder") |
| S17 | TI (PTSD) OR AB (PTSD) |
| S16 | MH ("stress disorders, post-traumatic+") |
| S15 | TI ("BD") OR AB ("BD") |
| S14 | TI (bipolar) OR AB (bipolar) |
| S13 | TI (manic) OR AB (manic) |
| S12 | TI (hypomani*) OR AB (hypomani*) |
| S11 | TI ("manic episode*") OR AB ("manic episode*") |
| S10 | TI ("hypomanic episode*") OR AB ("hypomanic episode*") |
| S9 | TI ("serious mental illness*") OR AB ("serious mental illness*") |
| S8 | TI ("severe mental illness*") OR AB ("severe mental illness*") |
| S7 | TI ("bipolar depressi*") OR AB ("bipolar depressi*") |
| S6 | TI ("manic depressi*") OR AB ("manic depressi*") |
| S5 | TI (mania*) OR AB (mania*) |
| S4 | TI ("affective disorder*") OR AB ("affective disorder*") |
| S3 | TI ("bipolar disorder*") OR AB ("bipolar disorder*") |
| S2 | MH ("bipolar and related disorders+") |
| S1 | (MH "Mood Disorders+") |
